# Who is asking ChatGPT health questions? Analysis of a nationally representative Australian community sample

**DOI:** 10.1101/2024.10.13.24315426

**Authors:** Julie Ayre, Erin Cvejic, Kirsten J McCaffery

## Abstract

ChatGPT and other generative artificial intelligence (AI) tools have dramatically changed the health information environment. Discussions about their benefits and risks need clear evidence about who is using them and why. This nationally representative survey estimates that one in ten Australians asked ChatGPT a health question in the past 6 months (95%CI: 8.5%□11.4%), with higher use in key priority groups: people born in a non-English speaking country, who speak a non-English language at home, and who have low health literacy. As usage increases, we need to equip community with knowledge and skills to use these tools safely.

---

With the launch of ChatGPT in 2022, Australians now have direct, easy access to a generative artificial intelligence (AI) model that can answer almost any health question. Though ChatGPT has capacity to vastly increase access to highly tailored health information, its errors are also well-documented, particularly for its early models, and variation in accuracy depending on the task and topic (1). As such, generative AI tools pose yet another potential challenge to health services and clinicians, in an environment already overwhelmed by misinformation (2). Discussions about potential benefits and risks of this new technology for health equity, patient engagement and safety need clear evidence about who and how many people are using ChatGPT, and the kinds of health questions they ask. This study aimed to examine ChatGPT use for health purposes in Australia.

We used the June 2024 wave of the Life in Australia™(3) panel to estimate self-reported use of ChatGPT in a nationally representative Australian sample (see eMethods). After providing consent, participants completed questions about how often they used ChatGPT for health purposes in the past 6 months, and for which health tasks. Those aware of ChatGPT but who had not used it for health purposes were asked about their intentions to do so in the next 6 months. Self-reported health literacy(4) was collected, with demographic information available through existing panel data. Social disadvantage corresponded to national quintiles for the Index of Socio-economic Advantage and Disadvantage(5). Scores were weighted to Australian population benchmarks using propensity scores. Associations were assessed using simple logistic regression, with p-values < .05 considered statistically significant. Unless otherwise stated, results refer to the unweighted analyses. The study was approved by the University of Sydney Human Research Ethics Committee (2024/HE000247).

Of the 2,951 invited panellists, 2,034 took part (68.9%). Sample demographics approximated those of the general population (Table 1). Most had heard of ChatGPT (weighted 84.7%, 95% confidence interval (CI): 83.0%□86.3%), with only a small proportion who had not (weighted 15.3%, 95%CI: 13.7%□17.0%). One in ten reported using ChatGPT in the last 6 months to ask health questions (weighted 9.9%, 95%CI: 8.5%□11.4%). This was more likely for participants who were younger, lived in a capital city, born in a non-English speaking country, spoke a language other than English at home, or had limited/marginal health literacy (Table 1).

**Table 1.**
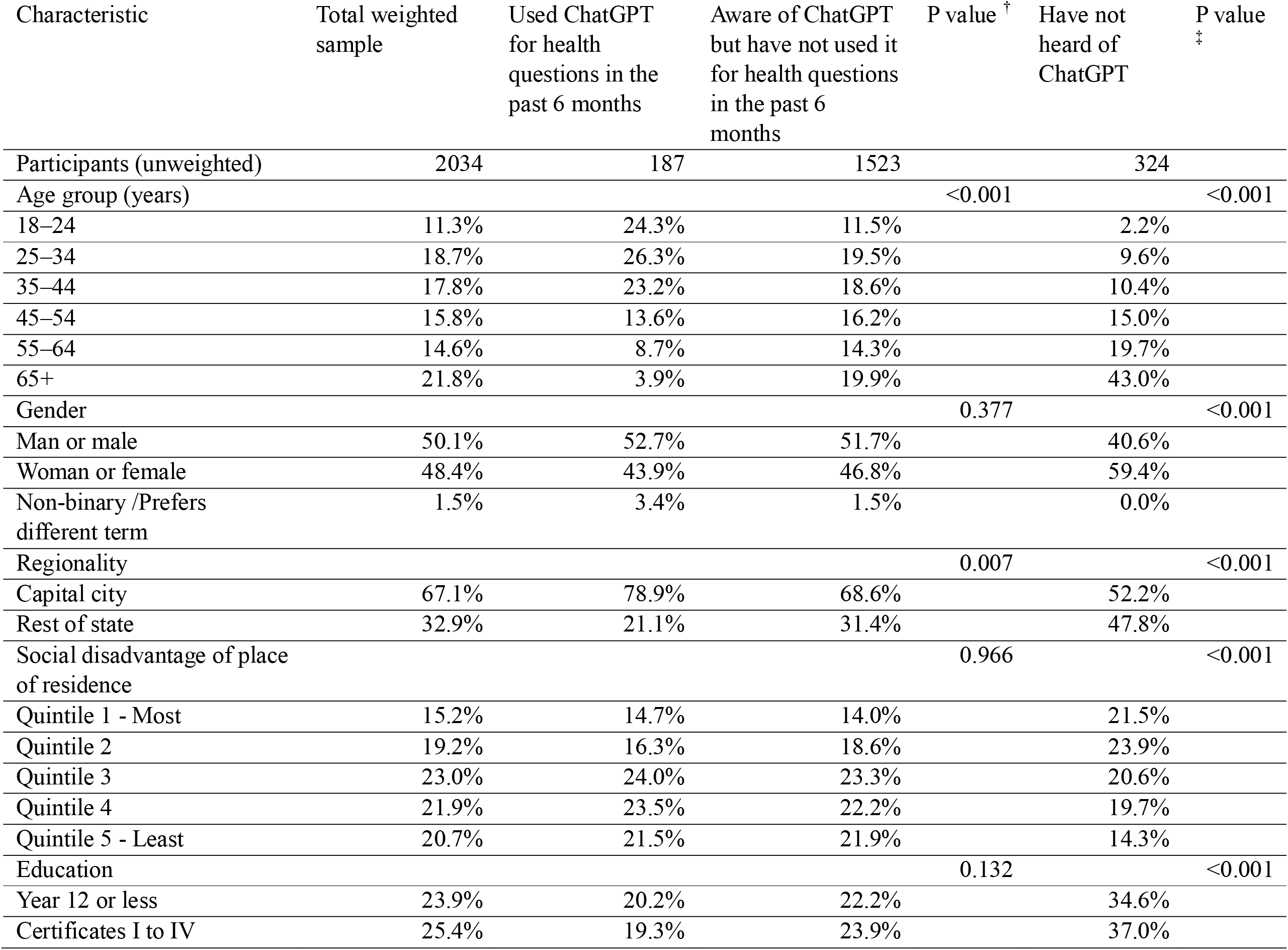
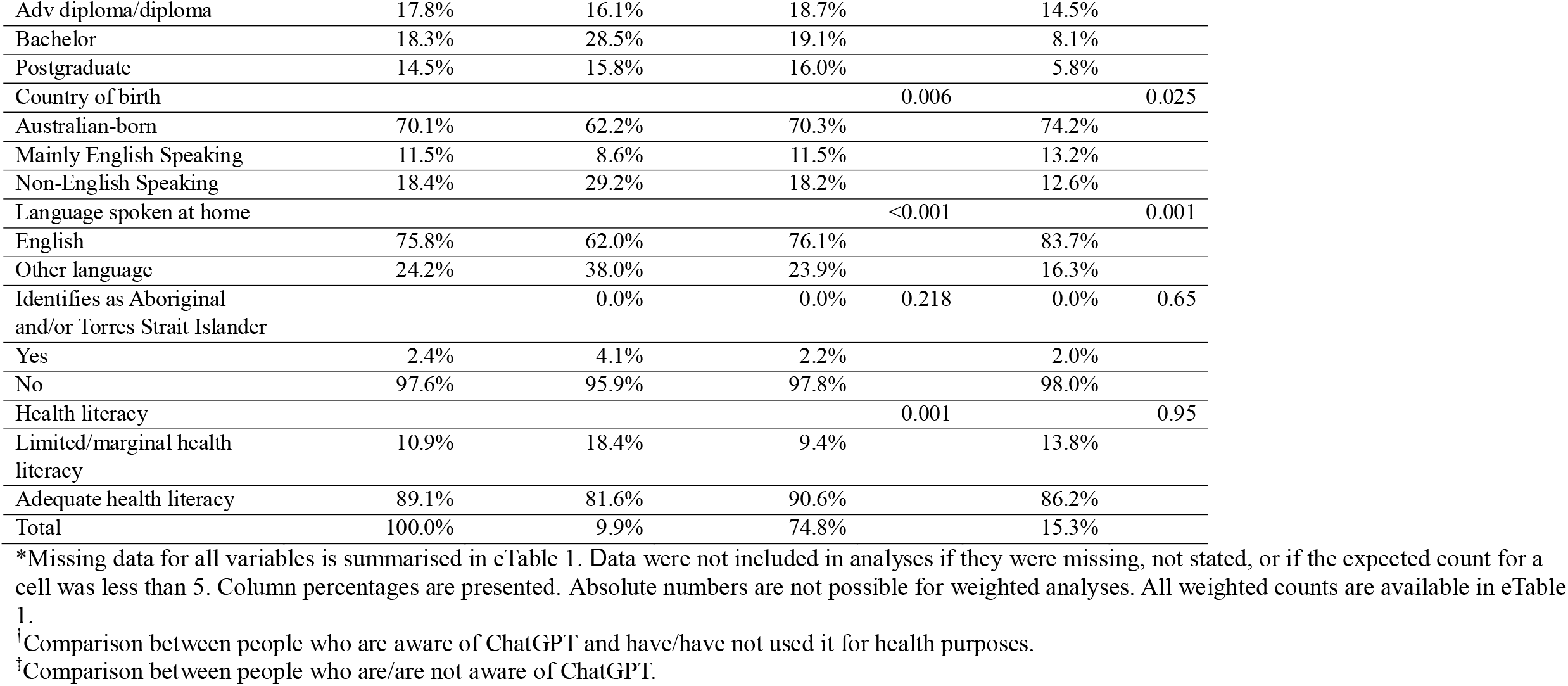
Participant characteristics (weighted to nationally representative population)*

Most people who used ChatGPT for health questions were fairly trusting of the tool (mean =4.39/5, standard deviation (SD)=0.88). The most common health questions related to learning about a specific health condition (89/187, 47.6%), finding what symptoms mean (70/187, 37.4%), finding out what actions to take (67/187, 35.8%), and understanding medical terms (65/187, 34.8%) (Table 2). Questions were categorised as ‘lower’ risk if they related to general descriptive health information, and ‘higher risk’ if they related to taking action that would typically require clinical advice. Three in five (115/187, 61.5%) reported asking at least one higher risk health question, and this was more likely amongst those born overseas in a mainly non-English speaking country compared to people born in Australia (odds ratio (OR)=2.62 95% CI:1.27□5.39), or who spoke another language at home compared to people who spoke English at home (OR=2.24, 95% CI: 1.16□4.32). No other associations were significant.

**Table 2.** Reported reasons for using ChatGPT for health in the past 6 months.

| **ChatGPT task\*** | **n (%)** |
| --- | --- |
| **Categorised as ‘higher risk’ questions** |  |
| Find out what my symptoms mean (or the symptoms of someone I know) | 70/187 (37.4%) |
| Find out what to do about a specific health issue that I or someone I know has | 67/187 (35.8%) |
| Find out if I or someone I know should see a doctor | 23/187 (12.3%) |
| Interpret results from blood tests or imaging | 16/187 (8.6%) |
| **Categorised as ‘lower risk’ questions** |  |
| Learn about a specific health condition | 89/187 (47.6%) |
| Understand medical terms | 65/187 (34.8%) |
| Learn more about a medicine, test or treatment (e.g. safety, side effects or interactions) | 57/187 (30.5%) |
| Learn about healthy lifestyles | 53/187 (28.3%) |
| Help create a plan to improve my health (or the health of someone I know) | 44/187 (23.5%) |

\*ChatGPT tasks were not mutually exclusive (more than 1 could be selected).

For those aware of ChatGPT but who had not yet used it for health purposes, 38.8% reported considering asking a health question in the next 6 months (591/1523). The most common tasks related to learning about a specific health condition (276/1523, 18.1%), understanding medical terms (256/1523, 16.8%), or finding out what symptoms mean (249/1523, 16.4%) (eTable 2). One quarter of these participants (375/1523, 24.6%) would consider asking at least one higher risk question, and this was more likely for participants with up to a Year 12 education (OR=1.76, 95%CI: 1.19□2.61) or an advanced diploma or diploma (OR=1.67, 95%CI: 1.11□2.51), compared to people with postgraduate education. No other associations were significant.

This exploratory study estimated that one in ten Australians asked ChatGPT a health question in the past 6 months, equivalent to almost 1.9 million adults(6). Given the sector’s explosive growth and the availability of similar tools (7), this is likely a conservative estimate of Australian health-related use of generative AI. This number is likely to grow: two in five people who had not yet asked ChatGPT a health question were considering doing so in the next 6 months. We also found health-related use of ChatGPT was higher for important priority groups who already face significant barriers to accessing care in Australia(8), including people born in a non-English speaking country, who do not speak English at home, and who have limited/marginal health literacy. More work is needed to understand which types of health questions pose a higher risk to community, acknowledging that this will change as the technology evolves. There is an urgent need to equip our community with new knowledge and skills to use generative AI tools safely, and to bring a strong equity lens to efforts that support this process.

## Data Availability

All data produced in the present study are available upon reasonable request to the authors

