## Supplementary Material for "Who is asking ChatGPT health questions? Analysis of a nationally representative Australian community sample"

### eMethods

Data were collected via the Life in Australia™ panel. This panel is made up of around 10,000 Australian adult residents (1, 2). Participants were initially randomly recruited via landline or mobile phone, and received a small financial incentive for joining the panel. Participants also receive a financial incentive for completing each subsequent survey. The panel uses probability-based sampling methods to cover both populations with and without internet access and participants can complete questions either online or by phone. The Life in Australia™ panel is Australia’s only probability-based online panel(3); all others are non-probability. Probability-based panels are considered significantly more accurate than non-probability-based panels (4). More specifically, comparative analyses suggest that the Life in Australia™ survey was demonstrably more accurate than four non-probability online panels, and obtained accuracy on par with telephone interviewing (3). In June 2024, Life in Australia™ launched a survey that contained the specific questions relevant to this study (eBox 1).

Invitation to take part (email or SMS) did not specifically describe the current study as it was one component of a larger survey. After opening the link, participants were then informed about this specific research project and consented to take part. They received up to 3 reminders to complete the survey. Participation was voluntary. Participants were required to complete all questions and did not review their answers at the end.

The survey was not pre-tested with participants, although bespoke survey items were based on co-designed items from another ChatGPT project. Items were developed with guidance from Life in Australia to ensure suitability for their panel surveys. The order of response options for each question was randomised (normal- or reverse-order).

Survey responses were anonymised prior to being made available to the research team. Two thousand and forty of the 2951 invited participants viewed the first page of the ChatGPT survey, 2036 saw the first question of the survey, and 2,034 completed the survey.

**eBox 1. ChatGPT survey items**

| **Survey item** | **Response options** |
| --- | --- |
| Q1. In the last 6 months how often have you used ChatGPT to answer questions about health? | Not at all  A few times  Once a month  Once a week  More than once a week  Never heard of ChatGPT  Not sure  Prefer not to say |
| Q2. *Participants who responded ≥ ‘a few times’ for Q1:*  In the last 6 months have you used ChatGPT to…  *All other participants:* In the next 6 months would you consider using ChatGPT to... | *Find out what my symptoms mean (or the symptoms of someone I know)  *Find out what to do about a specific health issue that I or someone I know has  Learn about a specific health condition  Learn about healthy lifestyles  Help create a plan to improve my health (or the health of someone I know)  Learn more about a medicine, test or treatment (e.g. safety, side effects or interactions)  *Find out if I or someone I know should see a doctor  Understand medical terms  *Interpret results from blood tests or imaging  Other  Not sure  Prefer not to say |
| Q3. How much do you trust what ChatGPT says? | Entirely  Quite a bit  Somewhat  A little bit  Not at all  Not sure  Prefer not to say |

*Categorised as higher risk health questions

*Self-reported use of ChatGPT for health questions*

Participants were categorised as using ChatGPT for health questions if they positively responded to Q1 and indicated that they had undertaken at least one health task in Q2. Participants who only selected the ‘Other’ category for Q2 were not considered using ChatGPT for health questions as pilot data showed that only a single instance of ‘other’ was related to health tasks. Participants who selected ‘Don’t know’ or ‘Never heard of ChatGPT’ were categorised as people who did not know what ChatGPT was. The remaining participants were categorised as people who were aware of ChatGPT but were not using it for health purposes.

ChatGPT health questions (Q2) were categorised as ‘lower’ risk if they related to general descriptive health information (e.g. what a medical term means) and ‘higher’ risk if they related to taking personalised advice or specific actions that would typically require advice from a clinician or health service (e.g. what treatment should the person use)(eBox 1). There is no current consensus about what kinds of health questions are more or less risky for community members to ask ChatGPT and other generative AI tools. Further, risk criteria are likely to shift as these technologies advance.

The validated single-item health literacy screener (5) was included, which categorises participants as low or high health literacy in response to the question “If you need to go to the doctor, clinic or hospital, how confident are you filling out medical forms by yourself?” Response options were extremely, quite a bit, somewhat, a little bit, or not at all. Participants responding ‘somewhat’ or less were considered to have limited or marginal health literacy. This self-report measure provides an estimate of health literacy, and is widely used in health literacy research, including in Australia (e.g. (6)) and internationally (e.g. (7)).

*Demographic items*

Demographic items were available as ‘panel variables’ and included age, gender, regionality corresponding to Greater Capital City Statistical Areas, social disadvantage corresponding to national quintiles for the Index of Socio-economic Advantage and Disadvantage)(8), highest level of education, Aboriginal and Torres Strait Islander status, using a language other than English at home, and country of birth for which ‘mainly English-speaking countries’ other than Australia included Canada, Ireland, New Zealand, South Africa, United Kingdom and United States of America. Further detail is available from the panel protocols (2).

### *Statistical analysis*

Scores were weighted to Australian population benchmarks using propensity scores. Benchmarks were weighted based on the National Health Survey 2020-21 (number of adults in household), and the Census 2021 updated using ABS Estimated Residential Population from July 2022 (age, highest educational attainment, gender, use a language other than English at home, capital city / rest of state, state/territory). Further detail about the weighting processes is available (2).

Data were analysed using SPSS Version 26. The SPSS Complex Samples function was used to obtain weighted estimates and corresponding confidence intervals. Differences in outcomes of interest across demographic variables were assessed using simple logistic regression. For these analyses data were not included if they were missing, not stated, and if the expected count for a cell was less than 5. P values <0.05 were considered statistically significant. Unless otherwise stated, descriptive and statistical reporting of results refers to the unweighted analysis.

### eTables

**eTable 1. Unweighted frequencies and weighted frequencies (and relative frequencies) of participant characteristics***

|  | **Unweighted** | | | | **Weighted** | | | |
| --- | --- | --- | --- | --- | --- | --- | --- | --- |
| **Characteristic** | **Total sample** | **Used ChatGPT for health questions in the past 6 months** | **Aware of ChatGPT but have not used it for health questions in the past 6 months** | **Have not heard of ChatGPT** | **Total sample** | **Used ChatGPT for health questions in the past 6 months** | **Aware of ChatGPT but have not used it for health questions in the past 6 months** | **Have not heard of ChatGPT** |
| Age group (years) |  |  |  |  |  |  |  |  |
| 18–24 | 182 | 37 | 138 | 7 | 230.82/2034 (11.3%) | 48.77/200.74 (24.3%) | 175.20/1522.43 (11.5%) | 6.85/1522.43 (2.2%) |
| 25–34 | 365 | 49 | 287 | 29 | 380.27/2034 (18.7%) | 52.84/200.74 (26.3%) | 297.47/1522.43 (19.5%) | 29.97/1522.43 (9.6%) |
| 35–44 | 362 | 42 | 289 | 31 | 362.13/2034 (17.8%) | 46.62/200.74 (23.2%) | 283.27/1522.43 (18.6%) | 32.23/1522.43 (10.4%) |
| 45–54 | 357 | 31 | 277 | 49 | 320.87/2034 (15.8%) | 27.20/200.74 (13.6%) | 246.99/1522.43 (16.2%) | 46.68/1522.43 (15.0%) |
| 55–64 | 324 | 19 | 235 | 70 | 296.10/2034 (14.6%) | 17.56/200.74 (8.7%) | 217.22/1522.43 (14.3%) | 61.32/1522.43 (19.7%) |
| 65+ | 444 | 9 | 297 | 138 | 443.81/2034 (21.8%) | 7.74/200.74 (3.9%) | 302.29/1522.43 (19.9%) | 133.78/1522.43 (43.0%) |
| Gender |  |  |  |  |  |  |  |  |
| Man or male | 958 | 100 | 743 | 115 | 1018.83/2034 (50.1%) | 105.73/200.74 (52.7%) | 786.90/1522.43 (51.7%) | 126.21/1522.43 (40.6%) |
| Woman or female | 1052 | 84 | 759 | 209 | 984.64/2034 (48.4%) | 88.10/200.74 (43.9%) | 711.91/1522.43 (46.8%) | 184.63/1522.43 (59.4%) |
| Non-binary /Prefers different term | 23 | 3 | 20 | 0 | 29.84/2034 (1.5%) | 6.91/200.74 (3.4%) | 22.93/1522.43 (1.5%) | 0.00/1522.43 (0.0%) |
| Refused | 1 | 0 | 1 | 0 | 0.69/2034 (0.0%) | 0.00/200.74 (0.0%) | 0.69/1522.43 (0.0%) | 0.00/1522.43 (0.0%) |
| Regionality |  |  |  |  |  |  |  |  |
| Capital city | 1392 | 146 | 1064 | 182 | 1360.43/2034 (66.9%) | 157.18/200.74 (78.3%) | 1041.32/1522.43 (68.4%) | 161.94/1522.43 (52.1%) |
| Rest of state | 635 | 40 | 454 | 141 | 666.07/2034 (32.7%) | 42.07/200.74 (21.0%) | 475.99/1522.43 (31.3%) | 148.01/1522.43 (47.6%) |
| Unable to establish | 7 | 1 | 5 | 1 | 7.50/2034 (0.4%) | 1.49/200.74 (0.7%) | 5.12/1522.43 (0.3%) | 0.88/1522.43 (0.3%) |
| Social disadvantage of place of residence |  |  |  |  |  |  |  |  |
| Quintile 1 - Most | 308 | 27 | 212 | 69 | 308.20/2034 (15.2%) | 29.25/200.74 (14.6%) | 212.43/1522.43 (14.0%) | 66.53/1522.43 (21.4%) |
| Quintile 2 | 380 | 29 | 274 | 77 | 389.60/2034 (19.2%) | 32.43/200.74 (16.2%) | 282.98/1522.43 (18.6%) | 74.19/1522.43 (23.9%) |
| Quintile 3 | 453 | 42 | 345 | 66 | 465.97/2034 (22.9%) | 47.84/200.74 (23.8%) | 354.17/1522.43 (23.3%) | 63.96/1522.43 (20.6%) |
| Quintile 4 | 449 | 46 | 340 | 63 | 444.00/2034 (21.8%) | 46.83/200.74 (23.3%) | 336.16/1522.43 (22.1%) | 61.01/1522.43 (19.6%) |
| Quintile 5 - Least | 437 | 42 | 347 | 48 | 418.75/2034 (20.6%) | 42.91/200.74 (21.4%) | 331.58/1522.43 (21.8%) | 44.26/1522.43 (14.2%) |
| Unable to establish | 7 | 1 | 5 | 1 | 7.50/2034 (0.4%) | 1.49/200.74 (0.7%) | 5.12/1522.43 (0.3%) | 0.88/1522.43 (0.3%) |
| Education |  |  |  |  |  |  |  |  |
| Year 12 or less | 505 | 41 | 351 | 113 | 480.82/2034 (23.6%) | 39.72/200.74 (19.8%) | 335.44/1522.43 (22.0%) | 105.66/1522.43 (34.0%) |
| Certificates I to IV | 523 | 40 | 370 | 113 | 511.10/2034 (25.1%) | 37.81/200.74 (18.8%) | 360.00/1522.43 (23.6%) | 113.29/1522.43 (36.4%) |
| Adv diploma/diploma | 367 | 32 | 286 | 49 | 358.58/2034 (17.6%) | 31.62/200.74 (15.8%) | 282.59/1522.43 (18.6%) | 44.37/1522.43 (14.3%) |
| Bachelor | 327 | 43 | 258 | 26 | 368.43/2034 (18.1%) | 55.97/200.74 (27.9%) | 287.64/1522.43 (18.9%) | 24.82/1522.43 (8.0%) |
| Postgraduate | 294 | 29 | 249 | 16 | 290.64/2034 (14.3%) | 31.03/200.74 (15.5%) | 241.96/1522.43 (15.9%) | 17.66/1522.43 (5.7%) |
| Unable to establish | 18 | 2 | 9 | 7 | 24.43/2034 (1.2%) | 4.58/200.74 (2.3%) | 14.81/1522.43 (1.0%) | 5.04/1522.43 (1.6%) |
| Country of birth |  |  |  |  |  |  |  |  |
| Australian-born | 1439 | 119 | 1083 | 237 | 1423.75/2034 (70.0%) | 124.77/200.74 (62.2%) | 1068.45/1522.43 (70.2%) | 230.53/1522.43 (74.2%) |
| Mainly English Speaking | 230 | 14 | 175 | 41 | 233.45/2034 (11.5%) | 17.34/200.74 (8.6%) | 175.12/1522.43 (11.5%) | 40.99/1522.43 (13.2%) |
| Non-English Speaking | 363 | 54 | 263 | 46 | 374.77/2034 (18.5%) | 58.63/200.74 (29.2%) | 276.82/1522.43 (18.2%) | 39.31/1522.43 (12.6%) |
| Unknown | 2 | 0 | 2 | 0 | 2.03/2034 (0.1%) | 0.00/200.74 (0.0%) | 2.03/1522.43 (0.1%) | 0.00/1522.43 (0.0%) |
| Language spoken at home |  |  |  |  |  |  |  |  |
| English | 1565 | 123 | 1180 | 262 | 1541.94/2034 (75.8%) | 124.52/200.74 (62.0%) | 1157.26/1522.43 (76.0%) | 260.16/1522.43 (83.7%) |
| Other language | 468 | 64 | 342 | 62 | 491.08/2034 (24.1%) | 76.21/200.74 (38.0%) | 364.20/1522.43 (23.9%) | 50.67/1522.43 (16.3%) |
| Refused | 1 | 0 | 1 | 0 | 0.98/2034 (0.0%) | 0.00/200.74 (0.0%) | 0.98/1522.43 (0.1%) | 0.00/1522.43 (0.0%) |
| Identifies as Aboriginal and/or Torres Strait Islander |  |  |  |  |  |  |  |  |
| Yes | 45 | 6 | 33 | 6 | 48.36/2034 (2.4%) | 8.02/200.74 (4.0%) | 34.15/1522.43 (2.2%) | 6.20/1522.43 (2.0%) |
| No | 1982 | 179 | 1486 | 317 | 1977.08/2034 (97.2%) | 189.88/200.74 (94.6%) | 1484.08/1522.43 (97.5%) | 303.13/1522.43 (97.5%) |
| Refused/ Don’t know | 7 | 2 | 4 | 1 | 8.56/2034 (0.4%) | 2.84/200.74 (1.4%) | 4.20/1522.43 (0.3%) | 1.51/1522.43 (0.5%) |
| Health literacy |  |  |  |  |  |  |  |  |
| Limited/ marginal health literacy | 216 | 33 | 137 | 46 | 222.10/2034 (10.9%) | 36.87/200.74 (18.4%) | 142.59/1522.43 (9.4%) | 42.64/1522.43 (13.7%) |
| Adequate health literacy | 1813 | 154 | 1384 | 275 | 1808.21/2034 (88.9%) | 163.86/200.74 (81.6%) | 1378.28/1522.43 (90.5%) | 266.07/1522.43 (85.6%) |
| Don’t know/ did not answer | 5 | 0 | 2 | 3 | 3.69/2034 (0.2%) | 0.00/200.74 (0.0%) | 1.56/1522.43 (0.1%) | 2.13/1522.43 (0.7%) |
| **Total** | **2034** | **187** | **1523** | **324** | **2034/2034 (100.0%)** | **200.74/2034 (9.9%)** | **1522.43/2034 (74.8%)** | **310.81/2034 (15.3%)** |

*Relative frequencies (%) may not match those shown in Table 1 as analyses in Table 1 did not include data if they were missing, not stated, or if the expected count for a cell was less than 5. Column percentages are presented.

**eTable 2. Reported reasons for considering using ChatGPT for health in the next 6 months**

| **ChatGPT task*** | **n (%)** |
| --- | --- |
| **Higher risk** |  |
| Find out what my symptoms mean (or the symptoms of someone I know) | 249/1523 (16.3%) |
| Find out what to do about a specific health issue that I or someone I know has | 176/1523 (11.6%) |
| Interpret results from blood tests or imaging | 172/1523 (11.3%) |
| Find out if I or someone I know should see a doctor | 117/1523 (7.7%) |
| **Lower risk** |  |
| Learn about a specific health condition | 276/1523 (18.1%) |
| Understand medical terms | 256/1523 (16.8%) |
| Learn more about a medicine, test or treatment (e.g. safety, side effects or interactions) | 201/1523 (13.2%) |
| Learn about healthy lifestyles | 154/1523 (10.1%) |
| Help create a plan to improve my health (or the health of someone I know) | 132/1523 (8.7%) |
| **Would not use ChatGPT for health questions** | 932/1523 (61.2%) |

*ChatGPT tasks were not mutually exclusive (more than 1 could be selected).
